# Maternal respiratory syncytial virus (RSV) vaccine perceptions amongst pregnant women and mothers of infants and toddlers in England: a qualitative study

**DOI:** 10.64898/2026.03.27.26349494

**Authors:** Sadie Bell, Tracey Chantler, Alyson Passanante, Joseph Pryce, Kate Bisset, Louise Letley, Helen Campbell, Pauline Paterson

## Abstract

**Background:** Since September 2024, respiratory syncytial virus (RSV) vaccination has been available in England for pregnant women (from 28 weeks’ gestation).

**Aim:** To explore RSV knowledge and awareness, RSV vaccination perceptions and acceptability, and preferences for maternal vaccine delivery and communication amongst pregnant women and mothers of infants and toddlers in England.

**Methods:** Between July and November 2024, semi-structured qualitative interviews were performed with 30 mothers (youngest child under 2 years), two of whom were pregnant with a subsequent child. The study was conducted as a follow-on to a UK Health Security Agency survey of attitudes towards RSV vaccination amongst pregnant and post-partum women in England.

**Findings:** Although most mothers had heard of RSV, mothers with experience in health roles were more likely to understand the potential severity of RSV in infants. Likelihood of maternal RSV acceptance was reported as high, with most mothers considering RSV vaccination as beneficial in protecting infants.

Most mothers preferred a hybrid approach to vaccine communication, with information available online (e.g. through the NHS website), via written sources (e.g. NHS produced leaflet), and through talking with midwives. For convenience, most mothers preferred the option of fitting vaccinations within the antenatal midwifery appointment schedule rather than going to general practice for a separate appointment.

**Conclusion:** To support maternal RSV vaccination decision-making and access, women need vaccine information early in pregnancy; information provision through a range of different sources (i.e. online, paper, in-person); and vaccination delivery in a convenient location (i.e. as part of antenatal appointments).

## Introduction

Respiratory syncytial virus (RSV) is a highly infectious virus and a common cause of respiratory tract infections in humans (UK Health Security Agency (UKHSA), 2025a). Although RSV infections can occur at any time of the year, infections are more common in colder months (October to March) (UKHSA, 2025a). RSV usually causes mild self-limiting illness with symptoms comparable to a common cold, but symptoms can be severe in older adults and infants who are more likely to develop lower respiratory tract infections (UKHSA, 2025a). In infants, RSV can cause bronchiolitis which can result in breathing difficulties, potentially requiring hospitalisation (Joint Committee on Vaccination and Immunisation (JCVI), 2023). Prior to RSV vaccination roll-out, there were an estimated 33,500 RSV-associated annual UK hospital admissions in children <5 years (Reeves et al., 2017) and 20 to 30 RSV-associated infant deaths (Cromer, et al., 2017).

Since 1^st^ September 2024, RSV vaccination is recommended routinely in pregnancy (from 28 weeks’ gestation) in England. Through transplacental transfer of maternal antibody, maternal vaccination helps protect infants for the first six months of life, when they are at greatest risk of severe RSV infection. Since being introduced in England and Scotland (in August 2024), RSV vaccination has been found to be highly effective (72%) in preventing infant hospitalisations when mothers were vaccinated >14 days before childbirth (Williams et al., 2025). The vaccine is free at the point of delivery for pregnant women and provided by maternity services with a small proportion of women vaccinated through general practice (GP) (UKHSA, 2025a).

RSV vaccination is one of three vaccines currently recommended during pregnancy. Seasonal influenza vaccination is recommended at any stage of pregnancy, generally between September and the end of March. Pertussis (Whooping cough) vaccination is offered around 20 weeks’ gestation (coinciding with mid-pregnancy ultrasound scans) but can be given anytime between 16 and 32 weeks’ gestation. As of spring 2025, pregnant women are no longer routinely eligible for COVID-19 vaccination, unless they are immunosuppressed. The risk of hospitalisation and severe COVID-19 complications significantly declined during the Omicron wave of COVID-19 infection and ushered this change in early 2025 (Department of Health & Social Care, 2024).

We conducted a qualitative interview study to explore the acceptability of RSV vaccination in pregnancy before and in the early stages of RSV vaccine introduction in England, to help inform the maternal RSV vaccination implementation strategy. This study was a follow-on to a June-July 2023 UK Health Security Agency (UKHSA) survey of 1061 pregnant and post-partum (up to 6 months post birth) women living in England (Broad et al., 2025). The survey determined whether respondents viewed RSV as serious for new babies, and their likelihood of accepting maternal RSV vaccination. The results indicated that 89.5% of respondents would be likely or highly likely to accept maternal RSV vaccination (Broad et al., 2025); in line with similar survey findings (Fitzpatrick et al., 2026; Paulson et al., 2025). Although maternal RSV vaccine acceptability was high in the UKHSA survey, there was some evidence of lower acceptability amongst mothers in minority ethnic groups (Broad et al., 2025). Our study aimed to gain deeper insights into maternal RSV awareness, maternal RSV vaccination acceptability, and preferences for maternal vaccine delivery and communication.

## Methods

### Recruitment

After completing the UKHSA survey, respondents were asked to provide their contact details (email and/or phone number) if they were interested in taking part in further RSV-related research. Respondents who provided contact details were purposively selected based on a range of characteristics including ethnicity, self-reported disability and geographical location. In our sampling, we sought to maximise diversity and achieve representation across these characteristics, particularly ethnicity, to explore the association between ethnicity and maternal RSV vaccination acceptability. In the preliminary UKHSA survey analysis, a potential association between disability status and RSV acceptance was indicated. Although this was not found in the final survey analysis, in planning our study we aimed to achieve representation amongst participants by self-reported disability.

We contacted women who reported they would or would not be likely to accept maternal RSV vaccination in a current or hypothetical future pregnancy, to explore the nuances of their reasons for accepting or declining maternal RSV vaccination.

Sampled participants were emailed an information sheet, detailing the study objectives and explaining all aspects of participation, including the right to withdraw from the research. Written informed consent was obtained from each participant.

### Data collection

Interviews were conducted one-to-one by TC, JP, PP and KB. Interviewers JP and KB were trained to conduct interviews by TC and PP, researchers with extensive experience in qualitative methods and the research topic. Interviews lasted approximately 30 minutes each and were conducted via Zoom or telephone call. All interviews were audio-recorded and reflective notes were taken during the interaction. A topic guide, developed drawing on the UKHSA survey results, was used to assist the interviews. This included questions on views on vaccinations, vaccinations received during pregnancy, awareness of maternal RSV, and preferences around vaccine delivery and communication.

Towards the mid-point of the interview, interviewers showed participants a copy of an NHS information leaflet on maternal RSV vaccination. After reviewing this information, interviewees were asked about acceptance of a maternal RSV vaccine in their current or a hypothetical future pregnancy.

Interview participants received a £20 gift voucher as a thank you for their time and contribution.

### Data analysis

Interviews were transcribed verbatim and analysed thematically in NVivo (Release 1.6.1) using the stages outlined by Braun and Clarke (2006): data familiarisation, coding and theme identification and refinement. Data analysis was conducted by AP and SB, who coded the same 5 transcripts as a test of reliability. A coding frame was developed by AP and SB, which was updated when new codes were identified. To enhance the rigour of the analysis, coding approaches and data interpretations were discussed by SB, AP, TC, JP and PP.

## Findings

### Participants

Almost one third (29.9% *n*=317) of UKHSA survey respondents provided their contact details. Most respondents reported they would be likely to accept a maternal RSV vaccination (92.1%; *n*=292). In total, 123 women were sent an email invitation. Of these, 30 took part in an interview, 84 did not respond, 6 responded initially but did not follow through with an interview, and 3 declined to participate.

All UKHSA survey respondents from minority ethnic groups who agreed to be contacted were invited to interview (n=40). Of these, 12 participated who self-reported as: other Mixed/multiple ethnic background (n=3); other White background (n=3); Black, Black British, Caribbean or African (n=2); Asian/Asian British Indian (n=2); Asian/Asian British Chinese (n=1); and White and Black Caribbean (n=1). The remaining 18 participants self-reported as White (English, Welsh, Scottish, Northern Irish, and British).

Participants were aged 25-34 years (n=15), 35-44 years (n=14), and 18-24 years (n=1)

When asked in the UKHSA survey if they experienced any long-term health problems or disabilities (i.e. which had lasted/were expected to last at least 12 months), that limited their day-to-day activities, most interview participants reported ‘not at all’ (n=20), followed by ‘limited a little’ (n=9) and ‘limited a lot’ (n=1).

Participants lived in the following regions of England: South East (n=7); Greater London (n=5); West Midlands (n=5); South West (n=3); East Midlands (n=3); East of England (n=2); North West (n=2); Yorkshire and Humber (n=2); and the North East (n=1).

Most participants were first time mothers (n=26), two participants were pregnant at interview and already had children, and one participant had given birth to twins. The age of the participants’ youngest child at interview ranged from 7 to 21 months

**Table 1:** Overview of participant characteristics.

| Participant Number | Age (years) | Ethnicity | Age of youngest child | Number of children |
| --- | --- | --- | --- | --- |
| 1 | 25-34 | White, English/ Welsh/ Scottish/ Northern Irish/ British | 0-12 months | 1 |
| 2 | 35-44 | Asian/ Asian British, Chinese | 0-12 months | 1 |
| 3 | 18-24 | White, English/ Welsh/ Scottish/ Northern Irish/ British | 12+ months | 2 or more |
| 4 | 25-34 | White, English/ Welsh/ Scottish/ Northern Irish/ British | 12+ months | 1 |
| 5 | 35-44 | White, English/ Welsh/ Scottish/ Northern Irish/ British | 0-12 months | 1 |
| 6 | 25-34 | White, English/ Welsh/ Scottish/ Northern Irish/ British | 0-12 months | 1 |
| 7 | 25-34 | Any other white background | 12+ months | 1 |
| 8 | 25-34 | White, English/ Welsh/ Scottish/ Northern Irish/ British | 0-12 months | 1 |
| 9 | 35-44 | Black/ African/Caribbean/Black British, African | 12+ months | 2 or more |
| 10 | 35-44 | Any other white background | 12+ months | 1 |
| 11 | 25-34 | White, English/ Welsh/ Scottish/ Northern Irish/ British | 0-12 months | 1 |
| 12 | 35-44 | Black/ African/Caribbean/Black British, African | 12+ months | 1 |
| 13 | 35-44 | White, English/ Welsh/ Scottish/ Northern Irish/ British | 12+ months | 1 |
| 14 | 35-44 | White, English/ Welsh/ Scottish/ Northern Irish/ British | 0-12 months | 2 or more |
| 15 | 35-44 | White, English/ Welsh/ Scottish/ Northern Irish/ British | Missing data | 1 |
| 16 | 25-34 | Any other white background | 0-12 months | 1 |
| 17 | 25-34 | White, English/ Welsh/ Scottish/ Northern Irish/ British | 0-12 months | 1 |
| 18 | 35-44 | White, English/ Welsh/ Scottish/ Northern Irish/ British | Missing data | 1 |
| 19 | 25-34 | White, English/ Welsh/ Scottish/ Northern Irish/ British | 12+ months | 1 |
| 20 | 35-44 | White, English/ Welsh/ Scottish/ Northern Irish/ British | 12+ months | 2 or more |
| 21 | 25-34 | White, English/ Welsh/ Scottish/ Northern Irish/ British | 12+ months | 2 or more |
| 22 | 35-44 | Any other Mixed/ multiple ethnic background | 0-12 months | 1 |
| 23 | 25-34 | White, English/ Welsh/ Scottish/ Northern Irish/ British | 12+ months | 1 |
| 24 | 35-44 | Any other Mixed/ multiple ethnic background | 12+ months | 1 |
| 25 | 25-34 | Asian/ Asian British, Indian | 12+ months | 1 |
| 26 | 25-34 | White, English/ Welsh/ Scottish/ Northern Irish/ British | 0-12 months | 2 or more |
| 27 | 35-44 | Asian/ Asian British, Indian | 12+ months | 1 |
| 28 | 25-34 | Mixed/ multiple ethnic groups, White and Black Caribbean | 0-12 months | 1 |
| 29 | 35-44 | White, English/ Welsh/ Scottish/ Northern Irish/ British | 0-12 months | 1 |
| 30 | 25-34 | White, English/ Welsh/ Scottish/ Northern Irish/ British | 12+ months | 2 or more |

### Vaccination uptake during recent pregnancy

During their most recent pregnancy, although recall was difficult for some, most mothers reported receiving pertussis vaccination (n=26). Fewer mothers had received flu (n=17) and COVID-19 (n=9) vaccinations, with COVID-19 still a recommended vaccination during these women’s pregnancies. Only one mother reported receiving no maternal vaccinations. One mother was vaccinated in another country with a different vaccination schedule to the UK.

Some mothers reported not receiving flu vaccines as they were pregnant outside of flu vaccination season (n=2). Some had not been vaccinated against COVID-19 and flu during pregnancy as they had received a COVID-19 or flu vaccination in the months prior to pregnancy or had recently had flu or COVID-19 infection and considered themselves protected (n=4). One mother had declined the flu vaccination for medical reasons, and another because they were unwell when vaccination was offered.

Vaccination access was a barrier to vaccination for some, with some mothers reporting that they had not received the flu or pertussis vaccination because it was not offered or promoted by their midwives (n=2), or that flu vaccination was unavailable due to stock shortages (n=1).

Pertussis vaccination acceptance was high because mothers considered this vaccination very effective in protecting infants, which was their main motivation to vaccinate. Pertussis vaccines were trusted and considered safe, as vaccines that have ‘been around for ages’ and given to many mothers. Notably, most mothers perceived pertussis vaccination as much more important than flu and COVID-19 vaccination. This was primarily because they considered that pertussis vaccination is given for the direct benefit of the child, which was less commonly considered for flu and COVID vaccines, and that pertussis is particularly serious and therefore any minor vaccine side effects experienced by the mother due to pertussis vaccination would be acceptable.

*‘I think the moment you pee on the stick, and you realise that you are pregnant I think all your maternal instinct goes to the baby and protecting yourself sort of becomes second.’ (Participant #10)*

*‘The nurse at the doctor’s surgery would’ve told me it’s a way of trying to protect your baby in the first few weeks before they’re able to have their own vaccines for whooping cough and such. So that would’ve been probably the reason why I took it, that one over any of the vaccines that I refused.’ (Participant #20)*

Mothers who declined flu and COVID-19 vaccination during pregnancy reasoned that these vaccines were primarily for their protection, hence less important, particularly if they did not consider flu or COVID-19 as serious illnesses.

Mothers were least likely to have accepted the COVID-19 vaccine, with concerns raised about the relative newness and safety of this vaccine, and potential side effects. Reasons for flu vaccination decline included the view that flu is not particularly serious, low perceptions of personal risk, and concerns about potential side effects.

### Awareness of RSV

Most mothers had heard of RSV to some degree, some only because of participating in the UKHSA survey, but few understood what RSV is. Unsurprisingly, mothers that worked in jobs related to health had the greatest awareness and understanding of RSV.

Several mothers noted that they had vaguely heard about RSV through social media (notably TikTok and Instagram), including via messaging around not letting other people kiss their babies and through mother’s sharing their personal experiences of severe RSV infection in their child.

> *‘So, I think there were a couple of people on social media, like friends and family, who said, “It’s RSV season, make sure you don’t have random people kissing the baby.” That’s the first I’d heard of it, really.’ (Participant #22)*
>
> *‘You don’t get really taught about – yeah, you don’t get taught about. Although what I would say is TikTok, oh my god it literally inundated me with – and I hadn’t searched it I think. And all my NCT [National Childbirth Trust] friends were saying about RSV, this is what it looks like, and everyone videoing their children, “Oh this is my kid in hospital, look for these signs.” So social media actually has made it a lot more out there, I suppose, than it was previously I think.’ (Participant #2)*

Interestingly, social media was considered beneficial in raising awareness of RSV, although mothers expressed that they would not trust social media as a source for maternal vaccine information.

*‘…social media is good for making you aware that something exists. You know. That’s where I first heard about RSV, but I wouldn’t use it for any medical advice or listening to anybody saying, “This is why you should get the vaccine and this is why you shouldn’t get the vaccine.”‘ (Participant #13)*

Others had heard about RSV (or of bronchiolitis) through personal experience of their child, or a family member or friend’s child, experiencing complications from RSV. Another source of RSV awareness was through National Childbirth Trust (a charity that provides paid-for antenatal and postnatal courses) friends and other mothers. A few mothers had also heard about maternal RSV vaccination in the UK news, when RSV vaccination in pregnancy was made available.

All mothers reported hearing far less about RSV when compared to flu, COVID or whooping cough. This was unsurprising given that most of the participants had given birth before RSV vaccination in pregnancy had been introduced.

### Perceptions around RSV severity and burden

Amongst mothers that had heard of RSV, most considered that it could be severe.

*‘I know it can be fatal if it’s young babies. So, it seemed to me, just from the small amount that I read about it, that it’s quite similar almost to whooping cough, in the sense that it’s not really a problem for older people. But for babies it can be really serious.’ (Participant #30)*

*‘I think it can become quite serious, they can end up in intensive care.’ (Participant #28)*

One mother had heard about RSV from a relative living in another European country who was advised to keep her babies at home for *‘three months and keep all her other children at home from day care’* to protect them from RSV at a particularly prevalent time. This mother considered RSV *‘potentially very, very serious especially in the very young.’*

When interviewers discussed how prevalent RSV is, and the number of children hospitalised with RSV related-illnesses per year, mothers were generally less aware that RSV affects a high number of children, and affects them with varying degrees of severity.

*‘I didn’t realise that it affected most children but that some weren’t poorly, poorly, they just got colds. So, she could have had it, she has had many colds so she could have had it already and I wouldn’t know.’ (Participant #18)*

### Views on accepting a maternal RSV vaccination

Following the sharing of an NHS information leaflet about maternal RSV vaccination in their interview, mothers were asked for their views on accepting the vaccine. They generally held positive views towards accepting an RSV vaccination in pregnancy with the reasons for this including the view that vaccinating would protect their baby at a particularly vulnerable stage in their life, and that maternal RSV vaccination is safe.

*‘. it makes sense to take that vaccine because you are giv[ing] the baby the protection from you. The baby is building up immunity from you before it is born.’ (Participant #12)*

Of paramount importance was offering protection to the baby, not to themselves.

*‘I think I would, 100%. I just think if you can potentially protect your baby – not so much myself but the baby, I think – because imagine having the baby and then them getting it. I’d be like, what if I just got that and then I could’ve protected him?’ (Participant #21)*

Before accepting RSV vaccination in a future pregnancy, some mothers wanted more information about the vaccination, to reinforce or enhance their confidence in having it. More information was wanted on aspects including safety, any potential side effects, length of effectiveness, any need for boosters, any links with RSV vaccination development and COVID vaccines, whether the vaccination also offers protection to the mother, and what type of vaccine the RSV vaccine is (e.g. live or inactivated).

*‘I suppose I’d want to know what type of vaccine it is I suppose what, if anything, are the risks to the baby. And then I suppose that it’s going to protect you, how long the vaccine’s going to help protect the baby ‘ (Participant #1)*

Two mothers reported they would decline or most likely decline maternal RSV vaccination. One, who was pregnant at the time of interview, expressed concerns about the RSV vaccine which paralleled her concerns about the COVID-19 vaccine.

*‘I don’t feel that there’s enough long-term safety data on that at the moment to be 100% confident that there were not going to be any implications from taking a vaccine during pregnancy. I think that’s probably my main deciding factor. I think the – your baby vaccines, they’ve been out for years, years and years and they’ve got long, long-term safety data going back decades, even. Whereas this one, it’s quite a new thing and I don’t feel that the information is out there to prove it’s safe at the moment.’ (Participant #20)*

The other mother who reported she would likely decline maternal RSV vaccination highlighted she would need more information about the vaccine and indicated that she did not feel particularly at risk of RSV as she was taking precautions to stay in good health and protect her baby. These precautions - which she would also take in a hypothetical future pregnancy – included living a healthy and active lifestyle, limiting social contact in early infancy, and breastfeeding. It is important to note that both mothers had declined the COVID-19 and influenza vaccines, and one had also declined pertussis vaccination.

### Preferences for RSV vaccine information and communication sources

Most mothers discussed a preference for having a hybrid approach to vaccine communication, with information provided online, most notably through the NHS website, and through offline methods, such as via an NHS produced leaflet and talking with midwives.

*‘I think just because you get told so much information in all these midwife appointments, it is good to be told verbally but also take something back just to flick through. They tell you so much, some of it just does go over your head a little bit so it just gives you that chance to go back, read it if you want to then go on and like I probably would just go and look at it a bit more before agreeing to anything. It just gives you that opportunity.’ (Participant #17)*

It was important to participants for vaccine information to be received in early pregnancy, rather than just when vaccinations were due, to give them time to read the information and consider their decision.

One mother who had particular concerns about vaccinations and had declined most maternal vaccinations, was seeking for greater transparency in vaccine information and opportunities to talk to healthcare professionals about potential vaccine side effects.

*‘I think if things were a bit more upfront with*… *in the past when I’ve taken my children for vaccinations, no one has ever at any point said to me, “Actually there’s a small risk that your child could have any sort of complications from taking this” I think maybe if people were a little bit more open like that and did make you aware that actually there can be a very small chance that you can have a complication from taking this or that, I think it would make me a little bit more open to it.’ (Participant #20)*

Some mothers did their own additional vaccination research, using sources such as peer-reviewed studies, WebMD, the UKHSA, the World Health Organisation, charities supporting mothers such as “Pregnant Then Screwed” (Pregnant Then Screwed, 2026), and online pages reached through Google searches.

Midwives were widely regarded as trusted and well qualified healthcare professionals for mothers to talk to about vaccinations. In general, mothers had a good experience with their midwives, who were usually seen frequently during pregnancy. Only two participants mentioned seeing a health visitor during pregnancy.

Problems occurred when there was a lack of consistency in terms of seeing the same midwife at their appointments, and when appointments felt rushed. In these situation mothers felt there was not the opportunity to discuss maternal vaccinations.

*‘I definitely think it would be more reassuring if you had midwives who had the time to discuss these kinds of things with you.’ (Participant #30)*

Some mothers highlighted that they would discuss vaccinations with friends and family members, to get what they considered to be a less biased or vaccine invested perspective.

### Preferred maternal vaccination delivery settings

Convenience, in terms of time, location and ease of access, was the main priority for participants when considering where they would prefer to receive maternal vaccinations. For this reason, most participants reported that getting vaccinated in maternity settings (either in hospital or community locations, or in general practice if maternity appointments were here) and fitting vaccinations around existing antenatal appointments was preferable.

*‘*…*during pregnancy we have multiple visits with the midwife*… *So, I prefer if during the visit to antenatal clinic, if the vaccine is given, so that will be great, so I don’t have to go to the pharmacy or I don’t have to go separately to the GP clinic. So, during the visit if it has been arranged then that will be great.’ (Participant #25)*

One mother also highlighted that having pregnancy vaccinations in maternity settings reiterates that *‘you’re having it [the vaccine] because you’re pregnant and that’s the reason for having this’. (Participant #8)*

A few mothers considered vaccination at their general practice (outside of their maternity appointment) or pharmacy was easier because it was located near to their home or workplace.

*‘Anywhere that’s accessible*… *Like I got both the flu and the COVID [vaccine] in a pharmacy that I could just book*… *I haven’t really got a preference on where as long as the person is trained and accessible and easy to book, as well. Because obviously you’re still working for the majority of your pregnancy.’ (Participant #23)*

Others found that booking general practice appointments was difficult.

*‘I suppose at the antenatal clinic would probably be a bit easier to – because the GP, there was a bit of a faff going, so I had to go to the receptionist to make an appointment, so queue up to get the appointment. With the antenatal they would just do it by text and say, “Come in on Monday,” and that’s it. So, for ease for me, personally, it would have been the antenatal clinic.’ (Participant #22)*

Due to their focus on maternal health, several mothers voiced that they would prefer to be vaccinated by their midwife rather than by a different healthcare professional at their general practice. However, other mothers were happy for any professional with the right training to deliver maternal vaccinations.

*‘I think to me the vaccine bit is I suppose separate from a midwife. I’m happy for anybody to jab me in the arm, kind of thing. So, it’d probably be more of a convenience point for me rather than kind of who or where it was delivered*..*’ (Participant #13)*

## Discussion

Disease awareness, knowledge and perceptions are a key component in shaping maternal vaccine acceptability (Razai et al., 2024). Previous studies have reported varying levels of RSV awareness amongst pregnant women, with Wilcox et al reporting that amongst 314 pregnant women surveyed in the South of England, 88% had no/little awareness of RSV (2019). More recent UKHSA and England-wide research, which our study followed on from, found that most of the currently or recently pregnant women surveyed (931/1054, 88.3 %) perceived RSV as serious for babies (Broad et al., 2025).

In our qualitative study we found that most mothers had heard of RSV to some degree, some due to participating in the preceding UKHSA survey which contained a brief overview of RSV. Levels of awareness ranged from having just heard the name of the virus, to having a greater understanding of what RSV is, and what health conditions RSV infection can lead to and the potential severity of these in infants. Participants with backgrounds in health care and those with more comprehension of RSV and related research were more likely to consider RSV infections as serious for infants.

Our study indicated that social media (notably TikTok and Instagram) played a positive role in promoting RSV awareness amongst mothers, and that perhaps through parent stories, social media may also play a role in developing maternal vaccine awareness.

Most mothers reported that they would be likely to accept maternal RSV vaccination in a current or hypothetical future pregnancy; however, several mothers raised concerns about the newness of the vaccine and a lack of long-term safety data. These concerns echo those expressed about maternal COVID-19 vaccination amongst several mothers in our study, and has been previously reported (Skirrow et al., 2022), highlighting how the newness of a vaccine can undermine confidence in its safety. Other studies have also reported safety concerns, and fear of side effects, as reasons for potential maternal RSV vaccination decline (Drislane, Moore & Attwell., 2026; Paulson et al., 2025).

Of those mothers in our study who reported they would not accept RSV vaccination in a current or hypothetical future pregnancy, it is important to note that apart from the pertussis vaccine, they had declined maternal vaccinations in their most recent pregnancy. Maternal pertussis vaccination uptake has recently shown significant improvement, from 64.4% of women who gave birth in September 2024 being vaccinated, to 72.9% of women who gave birth in September 2025 (UKHSA, 2025b). This increase in uptake followed a large outbreak in 2024, with 14,894 confirmed cases and 11 infant deaths (UKHSA, 2025c) and highlights how disease visibility focuses perceptions of risk and influences vaccine-decision-making. Pertussis vaccine acceptance was higher than for other maternal vaccines in our sample, due to the direct infant protection that mothers attributed to this vaccine. They were less concerned about their own health than their unborn child and infants’ health and some stated they avoided maternal influenza vaccine for this reason. Messaging about all maternal vaccinations needs to emphasise how individual vaccines protect the infants and the mother’s health.

To address any vaccination concerns, and information needs, mothers considered that a hybrid approach (including physical leaflets and electronic sources) to RSV vaccine communication, and vaccine communication more broadly, would be most effective. In our study, several mothers would have liked additional information about RSV vaccination (e.g. whether the vaccination is live or inactivated), which may be specific to our participants, with almost a quarter of mothers having backgrounds in health or medicine. In this instance, due to the variety of mothers’ information needs, it is essential for vaccination providers to be able to signpost to scientific resources. The timing of maternal vaccine information is also important, with several mothers’ wanting information earlier in pregnancy, giving them more time to read and digest the information, and ask any questions prior to vaccination.

Importantly, mothers trusted vaccine recommendations provided by their midwife. When midwives actively endorsed vaccination and encouraged open discussions about vaccines, rather than simply handing over vaccination information to pregnant women, it had a positive impact in promoting vaccination. This finding is consistent with previous research, which has highlighted the vital role midwives have in promoting maternal vaccination (Razai et al., 2024), and the need for midwifes to be equipped and confident in answering vaccination questions and addressing concerns. Higher levels of maternal vaccination knowledge amongst healthcare professionals are also associated with healthcare professionals being more likely to recommend vaccination (Wilcox et al., 2019). We also found that women prefer to be vaccinated in the most convenient location to them, which during pregnancy was usually in maternity settings, with vaccination appointments slotted around antenatal check-ups.

The latest maternal RSV vaccination data indicated an increase in uptake, from 33.6% in September 2024 to 53.7% in June 2025 (UKHSA, 2025d; UKHSA, 2025e), however there are notable differences in uptake by ethnicity and geographical location (UKHSA, 2025e). In June 2025, the highest uptake was reported in the South-West commissioning region (62.6%), and the lowest was reported in the London commissioning region (46.3%) (UKHSA, 2025e), which mirrors coverage in other vaccination programmes. By ethnicity, in June 2025, the highest uptake was reported amongst the ‘Other ethnic group – Chinese’ category (68.4%) and the lowest uptake amongst the ‘Black or Black British -Caribbean’ category (31.3%). Future research should delve deeper into reasons for differences in uptake by geographical location and other sociodemographic factors such as ethnicity, maternal age, and parity.

### Strengths and limitations

To our knowledge, this is the first qualitative study to explore maternal respiratory syncytial virus vaccine perceptions and acceptability amongst mothers in England. A strength of this study was our ability to interview a diverse range of mothers, including mothers from ethnic minority groups and those reporting a disability or long-term health condition.

Limitations include that apart from 2, none of the other mothers were pregnant at the time of interview and therefore could only discuss their views on accepting maternal RSV vaccination during a future pregnancy. To address this limitation, we held two small public engagement events in a multi-cultural urban area with pregnant women in April and May 2025 to share our research findings. Our findings resonated with them, and they had similar views on RSV vaccine acceptability, and preferences for maternal vaccine communication and delivery.

Secondly, as only two participants stated they were unlikely to accept maternal RSV vaccination, we were unable to thoroughly explore reasons for non-acceptance. Gaining a deeper understanding from mothers about the concerns and information needs regarding RSV vaccination did however shed light on how to address potential barriers to acceptance.

## Conclusion

This study provides deeper insight into maternal RSV awareness, perceptions of maternal RSV vaccination, and preferences for vaccine communication and delivery in England. Awareness of RSV varied, and while most mothers indicated likely acceptance of maternal RSV vaccination, concerns about the vaccine’s newness and long-term safety influenced confidence. Perceived infant protection was an important driver of acceptability.

Mothers expressed diverse information needs, favouring a hybrid approach to communication delivered earlier in pregnancy. Midwives were identified as trusted and influential in decision-making, and convenient vaccination within maternity settings was preferred.

Overall, these findings highlight key factors shaping maternal RSV awareness, vaccine perceptions, and acceptability, alongside practical considerations for optimising maternal vaccine communication and delivery.

## Data Availability

All data produced in the present work are contained in the manuscript

## Acknowledgments

We would like to thank the mothers that took part in this study, we are very grateful for your time and contribution, without which this research would not be possible.

We would also like to thank Professor Helen Bedford (University College London) for her expertise and valuable comments and feedback on draft manuscripts.

## References

Braun, V. & Clarke, V. (2006). Using thematic analysis in psychology. Qualitative Research in Psychology. 3 (2) 77–101

Broad, J., et al. (2025). An England-wide survey on attitudes towards antenatal and infant immunisation against respiratory syncytial virus amongst pregnant and post-partum women. Vaccine. 62 [Online]. Available at: https://www.sciencedirect.com/science/article/pii/S0264410X25007790. [Accessed: 15/09/2025].

Cromer, D., et al. (2017). Burden of paediatric respiratory syncytial virus disease and potential effect of different immunisation strategies: a modelling and cost-effectiveness analysis for England. Lancet Public Health. 2(8):e367–e374. doi: 10.1016/S2468-2667(17)30103-2.

Drislane, S., Moore, H., & Attwell, K. Understanding parental decisions to decline or delay infant RSV immunisation, nirsevimab, in Western Australia in 2024. Vaccine.

Grills, L.A. & Wagner, A.L. (2023). The impact of the COVID-19 pandemic on parental vaccine hesitancy: a cross-sectional survey. Vaccine. 41 (41) 6127–6133.

Joint Committee on Vaccination and Immunisation (JCVI). (2023). Respiratory syncytial virus (RSV) immunisation programme for infants and older adults: JCVI full statement, 11 September 2023. [Online]. Available at:

Paulson, S., et al. (2025). Protecting Against Respiratory Syncytial Virus: An Online Questionnaire Study Exploring UK Parents’ Acceptability of Vaccination in Pregnancy or Monoclonal Antibody Administration for Infants. Pediatric Infectious Diseases Journal. 44 (2S) S158–S161. [Online]. Available at: doi: 10.1097/INF.0000000000004632. [Accessed: 25/01/2026].

Pregnant then Screwed. (2026). About Pregnant Then Screwed. [Online]. Available at: https://pregnantthenscrewed.com/about-maternity-discrimination/. [Accessed: 06/03/2026].

Public Health England. (2021). Surveillance of influenza and other seasonal respiratory viruses in the UK Winter 2020 to 2021. [Online]. Available at: https://webarchive.nationalarchives.gov.uk/ukgwa/20220511034044/https://assets.publishing.service.gov.uk/government/uploads/system/uploads/attachment_data/file/995284/Surveillance_of_influenza_and_other_seasonal_respiratory_viruses_in_the_UK_2020_to_2021-1.pdf. [Accessed: 07/11/2025].

Razai, M.S., et al. (2024). Facilitators and barriers to vaccination uptake in pregnancy: A qualitative systematic review. PLoS One. [Online]. Available at: 10.1371/journal.pone.0298407. [Accessed: 23/01/2026].

Reeves, R.M., et al. (2017). Estimating the burden of respiratory syncytial virus (RSV) on respiratory hospital admissions in children less than five years of age in England, 2007-2012. Influenza and Other Respiratory Viruses. 11 (2) 122–129.

Skirrow, H., et al. (2022). Women’s views on accepting COVID-19 vaccination during and after pregnancy, and for their babies: a multi-methods study in the UK. BMC Pregnancy and Childbirth. 22 (33). [Online]. Available at: 10.1186/s12884-021-04321-3. [Accessed: 22/01/2026]

UK Health Security Agency (UKHSA). (2025a) Respiratory syncytial virus: the green book, chapter 27a. Published 21 February 2025. [Online]. Available at: https://www.gov.uk/government/publications/respiratory-syncytial-virus-the-green-book-chapter-27a. [Accessed: 27/05/2025].

UKHSA. (2025b). Latest figures show strong uptake of whooping cough vaccine in pregnancy. [Online]. Available at: https://www.gov.uk/government/news/latest-figures-show-strong-uptake-of-whooping-cough-vaccine-in-pregnancy. [Accessed: 06/03/2026].

UKHSA. (2025c). Confirmed cases of pertussis in England by month, 2024. [Online]. Available at: https://www.gov.uk/government/publications/pertussis-epidemiology-in-england-2024/confirmed-cases-of-pertussis-in-england-by-month. [Accessed: 23/01/2026].

UKHSA. (2025d). RSV maternal vaccination coverage in England: September 2024. [Online]. Available at: https://www.gov.uk/government/publications/rsv-immunisation-for-older-adults-and-pregnant-women-vaccine-coverage-in-england/rsv-maternal-vaccination-coverage-in-england-september-2024. [Accessed: 04/09/2025].

UKHSA. (2025e). Respiratory syncytial virus (RSV) maternal vaccination coverage in England: April 2025. [Online]. Available at: https://www.gov.uk/government/publications/rsv-maternal-vaccination-coverage-in-england/respiratory-syncytial-virus-rsv-maternal-vaccination-coverage-in-england-april-2025#discussion. [Accessed: 04/09/2025].

Wilcox, C.R., et al. (2019). Attitudes of Pregnant Women and Healthcare Professionals Toward Clinical Trials and Routine Implementation of Antenatal Vaccination Against Respiratory Syncytial Virus: A Multicenter Questionnaire Study. Pediatric Infectious Diseases Journal. 38 (9) 944–951

Williams, T.C., et al. (2025). Bivalent prefusion F vaccination in pregnancy and respiratory syncytial virus hospitalisation in infants in the UK: results of a multicentre test-negative, case-control study. Lancet Child and Adolescent Health. 9 (9) 655–662.

